# Comparison of Minimum Inhibitory Concentration (MIC) as Measured by Etests and Agar Dilution in *Neisseria gonorrhoeae* Isolates Tested from 2018-2024

**DOI:** 10.1101/2025.07.27.25332272

**Authors:** Joshua A. Manuel, Jennifer J. Wenner, Michael E. DeWitt, Brinkley R. Bellotti, Cindy Toler, Elizabeth Palavecino, Candice J. McNeil

**Affiliations:** Section on Infectious Diseases, Department of Medicine, Wake Forest University School of Medicine, Winston-Salem, NC, USA; Department of Biology, Wake Forest University, Winston-Salem, NC, USA; Department of Pathology, Wake Forest University School of Medicine, Winston-Salem, NC, USA; North Carolina Strengthening the United States Response to Resistant Gonorrhoea (SURRG) Guilford County Department of Public Health, Greensboro, NC, USA

**Keywords:** *Neisseria gonorrhoeae*, Etest, Minimum Inhibitory Concentration, Antibiotic Resistance, Agar Dilution

## Abstract

**Background:** In the U.S. *Neisseria gonorrhoeae* is the second most common reportable STI amassing 601,319 cases in 2023. Notably, *N. gonorrhoeae* has developed antimicrobial resistance, necessitating ongoing surveillance. In response to this threat, the U.S. Centers for Disease Control and Prevention (CDC) has established antimicrobial resistance surveillance networks including Strengthening the U.S. Response to Resistant Gonorrhea (SURRG) and the Antibiotic Resistance Lab Network (ARLN). SURRG performs testing using a gradient strip method, the Etest, while ARLN labs perform testing by agar dilution, for determining MIC values for *N. gonorrhoeae*.

**Methods:** We compared the concordance of MIC values obtained using the Etest gradient strip method at a SURRG site in North Carolina compared to ARLN using the agar dilution method for three antibiotics: azithromycin, cefixime, and ceftriaxone.

The MIC values and the corresponding interpretations for each agent were analyzed according to the recommendations of the Clinical and Laboratory Standards Institute (CLSI). The essential agreements were assessed

**Results:** Between January 2018 and December 2024, a total of 1,951 *N. gonorrhoeae* had corresponding isolates from the ARLN lab available for comparison of which 1,892 had corresponding Etest and agar dilution results for all three antibiotics. We found high levels of agreement between both testing methods for each antimicrobial agent tested over six years including for strains isolated from distinct anatomical sampling sites.

**Conclusions:** The Etest method provides a robust alternative for accurately detecting antibiotic-resistant *N. gonorrhoeae* for public health surveillance, which may make them particularly useful in resource- or labor-limited settings.

**Short Summary:** A comparison of two antimicrobial susceptibility testing methods, agar dilution and Etest, for determining the MICs of several antimicrobial agents against Neisseria *gonorrhoeae* revealed high levels of concordance between the two testing methods.

## Introduction

*Neisseria gonorrhoeae* is the second most common reportable STI amassing 648,056 cases in 2023 [1]. *N. gonorrhoeae* antimicrobial resistance has steadily increased rendering multiple classes of antibiotics ineffective, leaving ceftriaxone, a third-generation cephalosporin, as the front-line treatment for *N. gonorrhoeae* [2–7]. This emergence of drug resistance has produced the critical need for robust surveillance and testing methodologies to monitor and combat the spread of antibiotic-resistant strains.

In response to growing concern surrounding antimicrobial resistance, the U.S. Centers for Disease Control and Prevention (CDC) and the World Health Organization (WHO) have established extensive antimicrobial resistance surveillance networks [8,9]. These networks play pivotal roles in monitoring the prevalence and spread of antibiotic resistance, identifying emerging resistance patterns, and informing public health interventions aimed at mitigating the spread of resistant strains. Despite these efforts, low- and middle-income countries are at higher risk for antimicrobial resistant pathogen spread in large part due to limited resource and labor allocation towards high quality surveillance [10,11].

The gold standard method for antimicrobial susceptibility testing and determination of minimum inhibitory concentration (MIC) in *N. gonorrhoeae* is via the agar dilution method. Though accurate, this method used mostly by reference laboratories is relatively complex and time consuming. It involves plating a standardized inoculum of bacteria on multiple agar plates, each containing antibiotics at doubling dilution concentrations. In contrast, the Etest gradient method (bioMerieux, Durham, NC) uses a plastic strip impregnated with a gradient of antimicrobial agents on a lawn of bacteria, offering a simpler and more practical alternative for determining MIC values. For this reason, the Etest and other gradient diffusion methods have become increasingly popular in clinical laboratories due to their ease of use and reduced labor requirements [12]. They may be especially beneficial in settings with a low volume of specimens for antimicrobial susceptibility testing. Many previous studies provide evidence that Etests are a reliable method for determining *N. gonorrhoeae* MIC values for several drugs [13–15], yet discordance between agar dilution and Etests has been reported, potentially due to differences in technique or reading expertise [16–18]. Establishing that there exists agreement between both tests is paramount given the use of both methods for surveillance.

In this study, we assess the concordance between the agar dilution and Etest methods in determining the *N. gonorrhoeae* MIC values of three antimicrobials azithromycin, cefixime, and ceftriaxone. Though azithromycin is no longer a component of *N. gonorrhoeae* treatment in the U.S, we have included it here to highlight historic trends. Using data from laboratories participating in two CDC-supported antibiotic resistance surveillance networks; the Antibiotic Resistance Lab Network (ARLN) and the Guilford County, North Carolina Public Health Department as part of the Strengthening the U.S. Response to Resistant Gonorrhea (SURRG) project [10], which employ agar dilution and Etests methods respectively, we characterize the concordance (essential agreement) in MIC values over time and by specimen source. We also analyze the ability of the Etest to measure MIC within the same susceptibility category of agar dilutions (categorical agreement). High levels of agreement between both testing methods confirms that the Etest method can be used or accurately detecting antibiotic-resistant *N. gonorrhoeae* for public health surveillance purposes, which may make them particularly useful in resource- or labor-limited settings.

## Materials and Methods

Specimens collected from the endocervix, pharynx, rectum, urethra and vagina as part of SURRG activities were submitted to the North Carolina SURRG laboratory for culture and susceptibility testing by Etest for azithromycin, cefixime, and ceftriaxone. *N. gonorrhoeae* organisms isolated from positive specimens were initially tested by Etest at the NC SURRG laboratory and then submitted for MIC confirmation by the agar dilution method at the CDC-assigned regional Antimicrobial Resistance Laboratory Network (ARLN) in Tennessee, which in addition to azithromycin, cefixime, and ceftriaxone, performs a broader susceptibility panel. Antimicrobial susceptibility testing (AST) data from January 2018 to December 2024 were compared for the two methods performed on the same organism to assess concordance of MIC results.

The testing methods slightly differ in the individual MIC concentration levels that are tested and reported and some of the lower dilutions tested were slightly different throughout the years Changes in Etest resolution, or in other words differences in minimum reportable MIC concentration values over time also led to discrepancy between recordable values. To better illustrate these differences, see Supplemental Tables 1-4 in Supplemental Digital Content 1 for all MIC pairs and the concentrations each lab reports for the three antibiotics with specifications on which MIC values are assumed to be at equivalent concentration levels.

Essential agreement of an MIC result was defined as an MIC within +/-1 dilution between the Etest compared to the reference MIC result obtained by agar dilution method. The CLSI M52 guideline for acceptable performance among clinical trial data for manufacturers states >=90% of testing results should be in essential agreement [19]. MIC value pairs on the same row of Supplemental Tables 1-4 or on two adjacent rows would be in essential agreement. Results are provided for exact MIC equivalency among pairs as well as results for pairs that are within ± 1 level (essential agreement). In Tables 2-9, negative dilution differences suggest a lower value recorded by the Etest as compared to agar dilution, whereas positive dilution differences suggest a higher MIC value recorded.

**Table 1.** Sample sizes and Spearman’s rank correlation for agar dilution and Etest pairs.

| Antibiotic | *n* pairs | Spearman's $\rho$ | p-value |
| --- | --- | --- | --- |
| Azithromycin | 1,893 | 0.767 | <0.001 |
| Cefixime | 1,901 | 0.522 | <0.001 |
| Ceftriaxone | 1,892 | 0.781 | <0.001 |

**Table 2.** MIC equivalency and essential agreement of azithromycin, cefixime and ceftriaxone.

| Antibiotic | N Pairs | Dilution Difference |  |  |  |  | Essential Agreement |
| --- | --- | --- | --- | --- | --- | --- | --- |
|  |  | ≤ -2 | -1 | 0 | 1 | ≥ 2 |  |
| Azithromycin | 1,893 | 69<br>(3.6%) | 434<br>(23%) | 1054<br>(56%) | 312<br>(16%) | 24<br>(1.3%) | 1,800<br>(95%) |
| Cefixime | 1,901 | 20<br>(1.1%) | 313<br>(16%) | 1482<br>(78%) | 80<br>(4.2%) | 6<br>(0.3%) | 1,875<br>(98%) |
| Ceftriaxone | 1,892 | 78<br>(4.1%) | 504<br>(27%) | 1141<br>(60%) | 150<br>(8.0%) | 19<br>(1.0%) | 1,795<br>(95%) |

Essential agreement for MIC results is reported as a percentage calculated by the number of all pairs that are considered in essential agreement (± 1 level) divided by the total number of tested pairs. Results are reported across the entire collection time and a yearly time series for each antibiotic; also, a subgroup analysis was performed to assess the results by site of the specimen source. Spearman’s rank correlation coefficient was used to further measure the association between the reported MIC’s ranked level. For MIC pairs not reported as in essential agreement, a two-sided two-sample proportion test with continuity correction was performed to assess if the SURRG lab tends to report lower or higher than the ARLN lab.

Designations for susceptibility categories and alerts for risk of resistance emergence for each sample were made using the 2020 CDC Treatment Guidelines for gonorrhea [20]. Cutoffs for alerts were MIC ≥0.125 μg/mL for ceftriaxone, ≥0.25 μg/mL for cefixime, and ≥2 μg/mL for azithromycin. Major disagreements between Etests and the agar dilutions were determined as pairs in which one MIC triggered an alert for potential emerging resistance by one method and the other method did not.

## Results

A total of 1,951 SURRG samples had corresponding samples from the ARLN lab available for comparison between January 2018 and December 2024, of which 1,852 had results for all three antibiotics. The remaining contained missing values for at least one lab result. The sample sizes for each can be seen in Table 1, along with the Spearman’s rank correlation coefficient of the ranked pairs, which is significant for all three antibiotics indicating a positive association between the SURRG and ARLN lab results.

### Essential Agreement

Overall, MIC essential agreement for azithromycin, cefixime, and ceftriaxone was 95%, 98%, and 95% respectively (Table 2). The SURRG lab has maintained >86% essential agreement each year for all three antibiotics compared. These yearly essential agreement rates can also be seen in Figure 1 alongside the yearly equivalence rates and in Tables 3-5; yearly essential agreement ranged from 91% to 98% for azithromycin, 96% to 99.7% for cefixime, and 86% to 99.5% for ceftriaxone.

**Figure 1.**
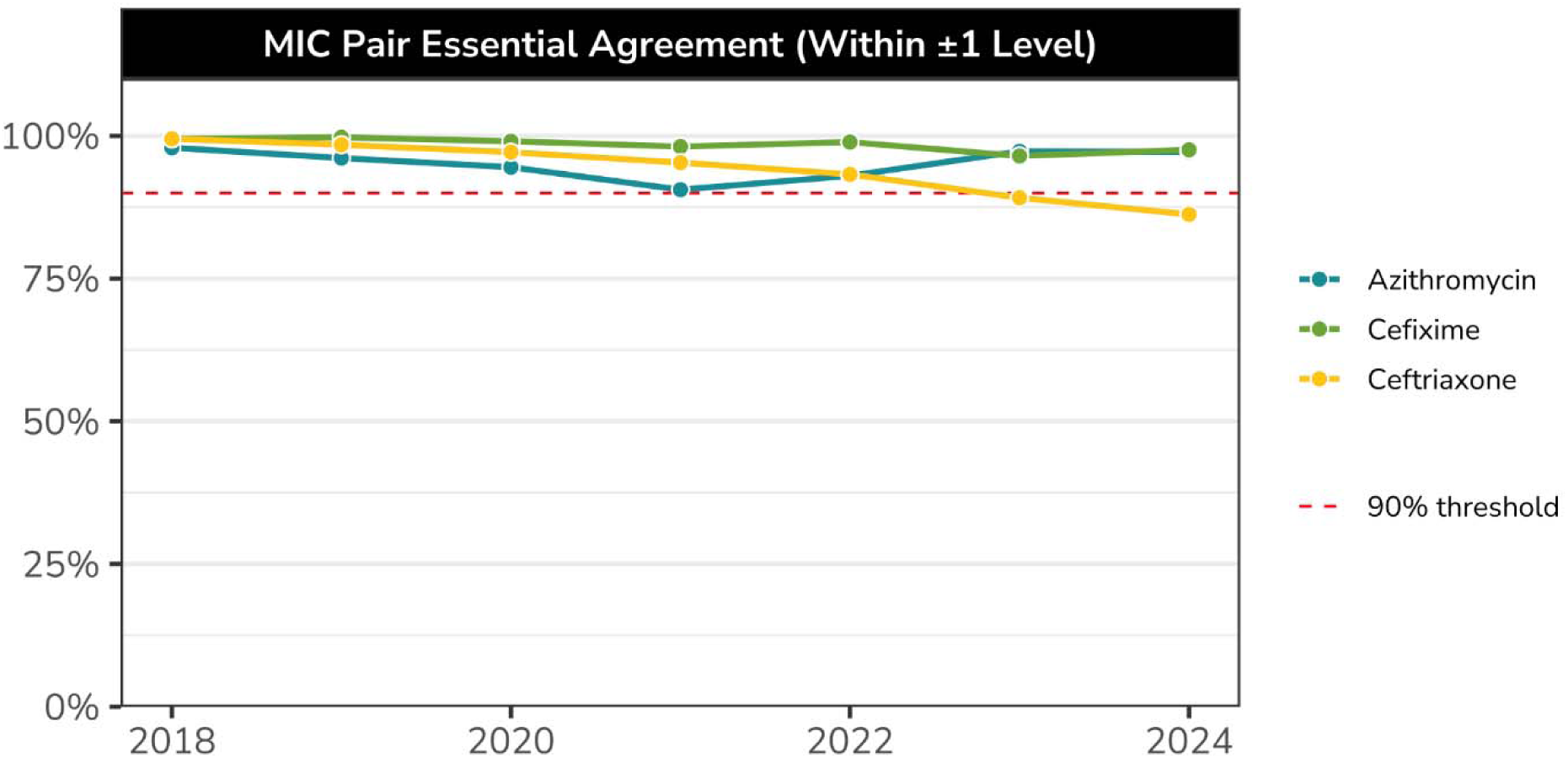
Yearly MIC pair equivalency (left) and concordance (right) rates from SURRG and ARLN lab results for azithromycin, cefixime, and ceftriaxone from 2018 to 2024.

**Table 3.**
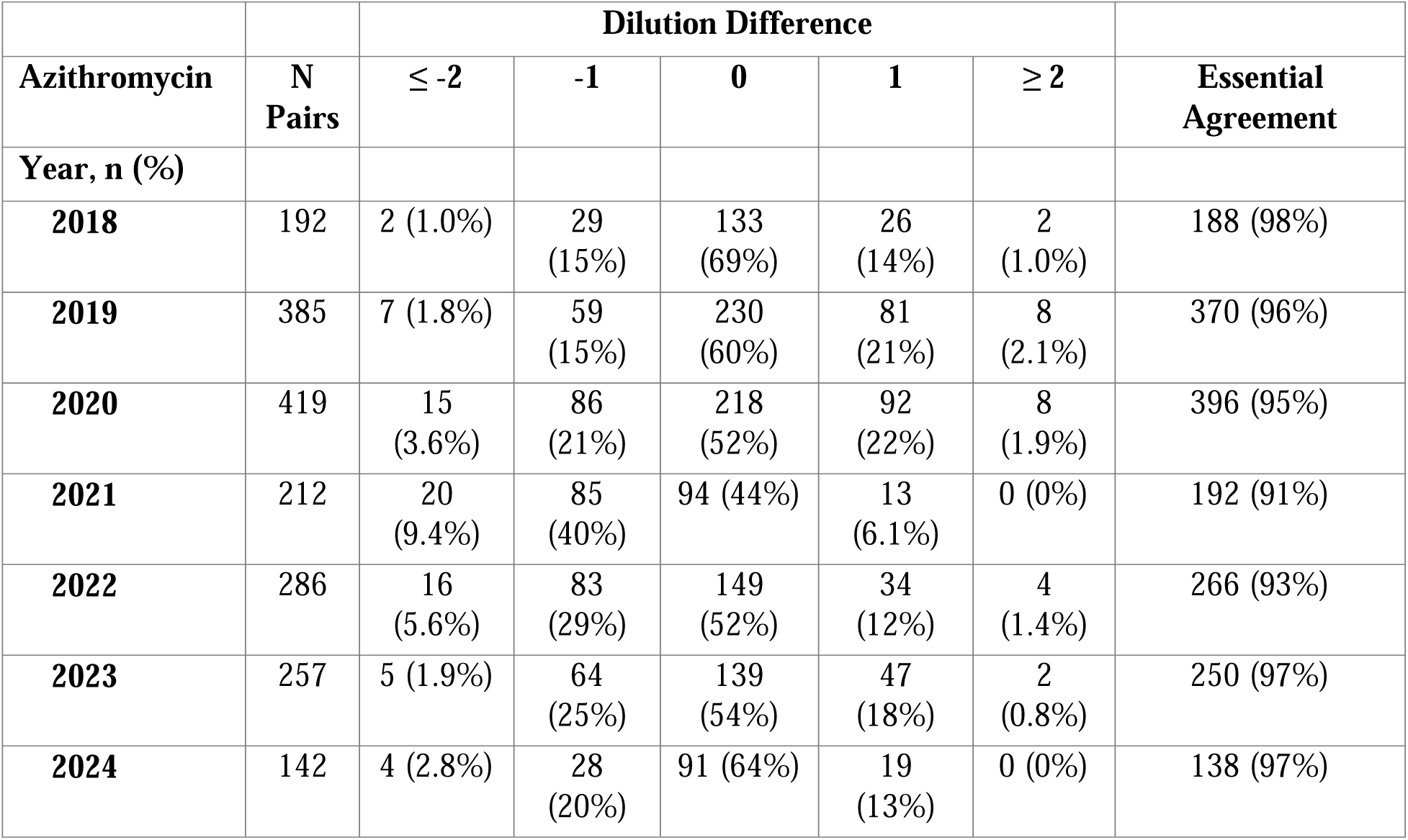
MIC equivalency and essential agreement of azithromycin, 2018-2023.

**Table 4.** MIC equivalency and essential agreement of cefixime, 2018-2023.

| Cefixime |  | Dilution Difference |  |  |  |  |  |
| --- | --- | --- | --- | --- | --- | --- | --- |
|  | N<br>Pairs | ≤ -2 | -1 | 0 | 1 | ≥ 2 | Essential<br>Agreement |
| Year, n<br>(%) |  |  |  |  |  |  |  |
| 2018 | 191 | 0<br>(0%) | 12<br>(6.3%) | 165<br>(86%) | 13<br>(6.8%) | 1 (0.5%) | 190 (99%) |
| 2019 | 385 | 0<br>(0%) | 50 (13%) | 316<br>(82%) | 18<br>(4.7%) | 1 (0.3%) | 384 (100%) |
| 2020 | 419 | 4 (1.0%) | 90 (21%) | 317<br>(76%) | 8 (1.9%) | 0 (0%) | 415 (99%) |
| 2021 | 213 | 2 (0.9%) | 24 (11%) | 174<br>(82%) | 11<br>(5.2%) | 2 (0.9%) | 209 (98%) |
| 2022 | 274 | 2 (0.7%) | 45 (16%) | 207<br>(76%) | 19<br>(6.9%) | 1 (0.4%) | 271 (99%) |
| 2023 | 255 | 9 (3.5%) | 71 (28%) | 173<br>(68%) | 2 (0.8%) | 0 (0%) | 246 (96%) |
| 2024 | 164 | 3 (1.8%) | 21 (13%) | 130<br>(79%) | 9 (5.5%) | 1 (0.6%) | 160 (98%) |

**Table 5.** MIC equivalency and essential agreement of ceftriaxone, 2018-2023.

| Ceftriaxone |  | Dilution Difference |  |  |  |  |  |
| --- | --- | --- | --- | --- | --- | --- | --- |
|  | N Pairs | ≤ -2 | -1 | 0 | 1 | ≥ 2 | Essential Agreement |
| Year, n (%) |  |  |  |  |  |  |  |
| 2018 | 192 | 1 (0.5%) | 6 (3.1%) | 184 (96%) | 1 (0.5%) | 0 (0%) | 191 (99%) |
| 2019 | 385 | 4 (1.0%) | 29 (7.5%) | 342 (89%) | 8 (2.1%) | 2 (0.5%) | 379 (98%) |
| 2020 | 419 | 9 (2.1%) | 117 (28%) | 254 (61%) | 36 (8.6%) | 3 (0.7%) | 407 (97%) |
| 2021 | 213 | 5 (2.3%) | 57 (27%) | 116 (54%) | 30 (14%) | 5 (2.3%) | 203 (95%) |
| 2022 | 267 | 17 (6.4%) | 102 (38%) | 116 (43%) | 31 (12%) | 1 (0.4%) | 249 (93%) |
| 2023 | 249 | 21 (8.4%) | 133 (53%) | 77 (31%) | 12 (4.8%) | 6 (2.4%) | 222 (89%) |
| 2024 | 167 | 21 (13%) | 60 (36%) | 52 (31%) | 32 (19%) | 2 (1.2%) | 144 (86%) |

For all three antibiotics, the Etest gave a lower MIC value more often than a higher MIC value MIC compared to the agar dilution method. 77%, 74% and 80% of all non-essential agreements were due to lower MIC readings by Etest for azithromycin, cefixime and ceftriaxone respectively (Table 6).

**Table 6.**
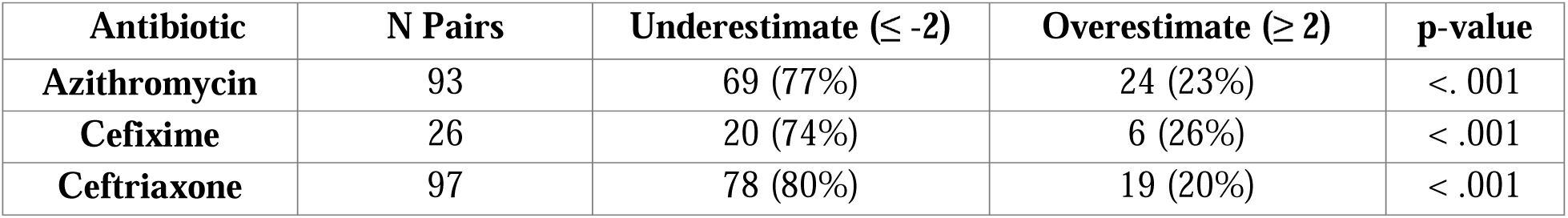
Total and Percentage of Underestimates and Overestimates Made by the Etest.

| Antibiotic | N Pairs | Underestimate (≤ -2) | Overestimate (≥ 2) | p-value |
| --- | --- | --- | --- | --- |
| Azithromycin | 93 | 69 (77%) | 24 (23%) | <. 001 |
| Cefixime | 26 | 20 (74%) | 6 (26%) | < .001 |
| Ceftriaxone | 97 | 78 (80%) | 19 (20%) | < .001 |

### Subgroup Analysis by Specimen Site

Specimens were collected from 6 different anatomical sites (Tables 7-9). For all antibiotics and organisms isolated from different specimen sites, the essential agreement was ≥ 93%.

**Table 7.** MIC equivalency and essential agreement of azithromycin by anatomical site.

| Azithromycin |  | Dilution Difference |  |  |  |  |  |
| --- | --- | --- | --- | --- | --- | --- | --- |
|  | N Pairs | ≤ -2 | -1 | 0 | 1 | ≥ 2 | Essential Agreement |
| Specimen Source, n (%) |  |  |  |  |  |  |  |
| Blood | 1 | 0 (0%) | 0 (0%) | 0 (0%) | 1 | 0 (0%) | 1 (100%) |
|  |  |  |  |  | (100%) |  |  |
| **Endocervical** | 171 | 7 (4.1%) | 39<br>(23%) | 85<br>(50%) | 39<br>(23%) | 1 (0.6%) | 163 (95%) |
| **Pharyngeal** | 314 | 11<br>(3.5%) | 76<br>(24%) | 173<br>(55%) | 50<br>(16%) | 4 (1.3%) | 299 (95%) |
| **Rectal** | 137 | 3 (2.2%) | 31<br>(23%) | 74<br>(54%) | 29<br>(21%) | 0 (0%) | 134 (98%) |
| **Urethral** | 1,123 | 44<br>(3.9%) | 257<br>(23%) | 644<br>(57%) | 162<br>(14%) | 16<br>(1.4%) | 1,063 (95%) |
| **Vaginal** | 147 | 4 (2.7%) | 31<br>(21%) | 78<br>(53%) | 31<br>(21%) | 3 (2.0%) | 140 (95%) |

### Susceptibility Category/Emerging Resistance Alert Analysis

The number of major disagreements in alert values, or in other words, the times in which the agar dilution method and Etests did not both alert for potential emerging resistance, were 49, 4 and 1 time for azithromycin, cefixime and ceftriaxone respectively (Table 10).

**Table 8.** MIC equivalency and essential agreement of cefixime by anatomical site.

| **Cefixime** |  | **Dilution Difference** |  |  |  |  |  |
| --- | --- | --- | --- | --- | --- | --- | --- |
|  | **N**<br>**Pairs** | **≤ -2** | **-1** | **0** | **1** | **≥ 2** | **Essential**<br>**Agreement** |
| **Specimen Source,**<br>**n (%)** |  |  |  |  |  |  |  |
| **Blood** | 1 | 0 (0%) | 1<br>(100%) | 0 (0%) | 0 (0%) | 0 (0%) | 1 (100%) |
| **Endocervical** | 169 | 1<br>(0.6%) | 25<br>(15%) | 130<br>(77%) | 11<br>(6.5%) | 2<br>(1.2%) | 166 (98%) |
| **Pharyngeal** | 319 | 4<br>(1.3%) | 63<br>(20%) | 234<br>(73%) | 17<br>(5.3%) | 1<br>(0.3%) | 314 (98%) |
| **Rectal** | 133 | 3<br>(2.3%) | 27<br>(20%) | 100<br>(75%) | 3<br>(2.3%) | 0 (0%) | 130 (98%) |
| **Urethral** | 1,133 | 11<br>(1.0%) | 180<br>(16%) | 902<br>(80%) | 37<br>(3.3%) | 3<br>(0.3%) | 1,119 (99%) |
| **Vaginal** | 146 | 1<br>(0.7%) | 17<br>(12%) | 116<br>(79%) | 12<br>(8.2%) | 0 (0%) | 145 (99%) |

**Table 9.** MIC equivalency and essential agreement of ceftriaxone by anatomical site.

| **Ceftriaxone** |  | **Dilution Difference** |  |  |  |  |  |
| --- | --- | --- | --- | --- | --- | --- | --- |
|  | **N**<br>**Pairs** | **≤ -2** | **-1** | **0** | **1** | **≥ 2** | **Essential**<br>**Agreement** |
| **Specimen Source,**<br>**n (%)** |  |  |  |  |  |  |  |
| **Blood** | 1 | 0 (0%) | 0 (0%) | 0 (0%) | 1 (100%) | 0 (0%) | 1 (100%) |
| **Endocervical** | 170 | 7<br>(4.1%) | 36 (21%) | 111<br>(65%) | 16<br>(9.4%) | 0 (0%) | 163 (96%) |
| **Pharyngeal** | 316 | 15<br>(4.7%) | 84 (27%) | 192<br>(61%) | 23<br>(7.3%) | 2 (0.6%) | 299 (95%) |
| **Rectal** | 131 | 7<br>(5.3%) | 46 (35%) | 66 (50%) | 10<br>(7.6%) | 2 (1.5%) | 122 (93%) |
| **Urethral** | 1,127 | 45<br>(4.0%) | 308<br>(27%) | 674<br>(60%) | 88<br>(7.8%) | 12<br>(1.1%) | 1,070 (95%) |
| **Vaginal** | 147 | 4<br>(2.7%) | 30 (20%) | 98 (67%) | 12<br>(8.2%) | 3 (2.0%) | 140 (95%) |

**Table 10.**
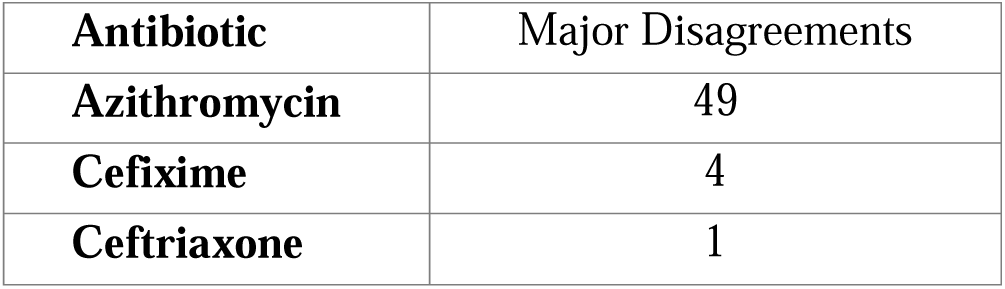
Number of major disagreements in alerts between the Etest and Agar Dilution.

## Discussion

Overall, we found that all three antimicrobials, with the exception of ceftriaxone in 2023 and 2024, achieved above the CLSI standard 90% level of essential agreement between agar dilutions and Etests across all years and inoculation sites for *Neisseria gonorrhoeae* MIC. Furthermore, major categorical disagreement for the preferred frontline antibiotic, ceftriaxone, occurred only once in nearly 2000 specimens tested. For MIC values that were not identical between the two methods, the Etest more often provided lower MICs. Importantly, these findings were consistent over a seven-year period, highlighting the potential utility of Etests for MIC determination in resource-limited settings. This approach may be especially valuable for detecting resistant *N. gonorrhoeae* strains and monitoring resistance trends in areas that might otherwise lack the capacity for agar dilution

Etest agreement remained consistent over time, even in the face of potential challenges such as staff turnover and variability in testing procedures or supplies. Notably, the downward trend in agreement for ceftriaxone, which was likely caused by the SURRG lab’s change to Etest strips with lower maximum and minimum antibiotic concentrations in late 2019, highlights the vulnerability of longitudinal MIC comparison to changes in methods. However, this disagreement also provides evidence for the robustness of Etests to provide accurate results, with the lowest level of agreement across all years still just below the FDA threshold at 86% essential agreement. The Etests robustness is further supported by its ability to avoid major categorical disagreements, or times when the Etest produces an MIC which disagrees with the agar dilutions MIC in such a way that the sample is incorrectly interpreted as susceptible or resistant, potentially changing drug choice for targeted therapy. For ceftriaxone, there was only one major disagreement out of the nearly 2,000 samples analyzed as well as similarly low rates of disagreement for azithromycin and cefixime. In other words, despite ceftriaxone falling below 90% agreement two years, the susceptibility category the Etest placed the sample in was identical to agar dilution in all but one specimen.

The Etest’s tendency to provide a lower MIC value compared to agar dilution is consistent with Liu et al. who previously described this finding in a similar study with only 105 *N. gonorrhoeae* isolates [16]. Unlike this study, however, our results cannot be explained by reading to the finer increments available on Etests. Though there is a subjective component to this test, results that seemed outside the range or unusual were always confirmed by a secondary reader. The SURRG lab reads and reports the MIC to the nearest doubling dilutions which match the antibiotic concentrations used in the standard agar dilutions per CDC reporting guidelines. Etests providing lower MIC values is seen across many drug classes and bacterial species [21–25] making it difficult to pinpoint an explanation for these results, but it seems more related to the Etest method rather than the organism tested.

Essential agreement across different anatomical sites is a crucial finding for confidence in Etests as an alternative for antibiotic resistance surveillance of *N. gonorrhoeae* isolated from different specimen types. Prioritizing speed and quickly accessible results in the context of public health surveillance has the potential to make MIC measurement more prone to causes for error. For example, specimen types harboring more extensive microbiomes, such as from the pharynx and rectum which are known to harbor other commensal *Neisseria* species [26], may coincide with contamination leading to difficulty in sample isolation for MIC measurement. Furthermore, heteroresistance, a phenomenon in which populations of bacteria contain minority-resistant subpopulations [27] has previously been described in *N. gonorrhoeae* [28], would cause discordance between Etests and agar dilutions due to the indeterminately large number of cells being tested on an Etest lawn, >10^7^ CFU and 10^4^ CFU respectively [29]. Different infection sites offer alternative selection pressures which may fuel greater genetic diversity amongst sampled populations. The environmental differences between anatomical sites include but are not limited to differences in opportunity for refuge from the immune system, variability in antibiotic concentration, nutrient availability and competition with other microbial populations, all of which provide unique growth opportunities and selection pressures that may result in the evolution of heteroresistance [30]. The high rates of concordance between the two tests further emphasize the strength of Etests to make accurate MIC determinations in spite of these potential causes for error.

Our results provide evidence that the Etest method offers a less labor-intensive alternative to agar dilution for antimicrobial susceptibility testing for *N. gonorrhoeae* and is suitable for use in a public health setting processing small volumes of specimens, which can potentially be leveraged in low-resource or low volume labs which require rapid results in individual specimens. The Etest method offers a robust alternative to the more laborious agar dilution method for surveillance of antibiotic-resistant *N. gonorrhoeae*.

## Supporting information

Supplemental Digital Content 1

## Data Availability

All data produced in the present study are available upon reasonable request to the authors

## Acknowledgements

We gratefully acknowledge the support and collaboration of the Centers for Disease Control and Prevention (CDC) Antimicrobial Resistance Laboratory Network (ARLN) and the North Carolina Strengthening the US Response to Resistant Gonorrhea (NC SURRG) network including our partners at the Centers for Disease Control and Prevention, the Guilford County Department of Public Health, the North Carolina Department of Health and Human Services, and Wake Forest University School of Medicine.

## Contributions

JM, JW, MD, BB data analysis, data interpretation, manuscript preparation and review. CJM, EP, and CT data interpretation, manuscript preparation and review. MD, JM, JW, BB and CJM had direct access to and verified the underlying data reported in the manuscript.

## Supplemental Digital Content

Supplemental Digital Content 1.docx

