## Supplemental Digital Content 1 for "Comparison of Minimum Inhibitory Concentration (MIC) as Measured by Etests and Agar Dilution in *Neisseria gonorrhoeae* Isolates Tested from 2018-2024"

Supplemental Table 1. Azithromycin SURRG and ARLN reported MICs

| Azithromycin MIC Equivalency | |
| --- | --- |
| SURRG | ARLN |
| 0.016 | 0.015 |
| 0.032 | 0.03 |
| 0.064 | 0.06 |
| 0.125 | 0.125 |
| 0.25 | 0.25 |
| 0.5 | 0.5 |
| 1 | 1 |
| 2 | 2 |
| 4 | 4 |
| 8 | 8 |

Supplemental Table 2. Cefixime SURRG and ARLN reported MICs

| Cefixime MIC Equivalency | |
| --- | --- |
| SURRG | ARLN |
| 0.016 | 0.002, 0.004, 0.008, 0.015 |
| 0.032 | 0.03 |
| 0.064 | 0.06 |
| 0.125 | 0.125 |
| 0.25 | 0.25 |

Supplemental Table 3. Ceftriaxone SURRG and ARLN reported MICs 2018-August 2019

| Ceftriaxone MIC Equivalency Sept 2019 - 2024 | |
| --- | --- |
| SURRG | ARLN |
| 0.016 | 0.001, 0.002, 0.004, 0.008, 0.015 |
| 0.032 | 0.03 |
| 0.064 | 0.06 |
| 0.125 | 0.125 |

Supplemental Table 4. Ceftriaxone SURGG and ARLN reported MICs Sept 2019-2023

| Ceftriaxone MIC Equivalency Sept 2019 - 2024 | |
| --- | --- |
| SURRG | ARLN |
| 0.002 | 0.001, 0.002 |
| 0.004 | 0.004 |
| 0.008 | 0.008 |
| 0.016 | 0.015 |
| 0.032 | 0.03 |
| 0.064 | 0.06 |
| 0.125 | 0.125 |
